# How does migratory status affect non-communicable disease risk in a global context? A systematic review

**DOI:** 10.1101/2021.08.31.21262947

**Authors:** Roy Gordon-Firing Sandberg

## Abstract

Challenges pertaining to the study of migrant health have been reported in medical and sociological literature. A literature review was thus conducted to gauge what research currently holds about the risk of non-communicable disease within migrant populations globally. The search strategy is outlined; CASP checklists were used to critically appraise articles, and the relevant data was synthesised and analysed. The research identifies several overarching quantitative themes regarding risk patterns. Recommendations are made.

## Introduction

The number of international migrants as a percentage of the global population sat at 3.4 in 2017, up from 2.8% in the year 2000. In absolute terms, this represented 258 million human beings. Taking into account internal migration (within their country of birth), the number reaches over 1/8th of the planet’s population (PAHO, 2018). From a public health perspective, particularly in high migrant turnover states such as the US or political unions such as the EU, the health outcomes of this subset of the population are extremely important in the long term. The diverse makeup of health systems (particularly public/private differentials), migratory trends, and other factors mean that an appropriate public health response needs both flexibility and a basis in scientific evidence.

Even though there are promising studies being undertaken, such as the HELIUS European cohort study, the literature indicates a gap in appropriate research output on the health of migrants, particularly on non-communicable diseases and maternal health (Sweileh, 2018). The existing research has found only minor differences in non-communicable disease (NCD) risk (Wang et al., 2019) for migrants as compared to non-migrants, but the very nature of a heterogeneous population such as that broadly defined as “migrants” implies that further research is warranted to identify any generalisable characteristics that could help governments and organisations design appropriate public health responses to emerging problems. Furthermore, a particular preventative approach regarding risk factors for NCD (Pheiffer, 2021) would certainly save many lives, not to mention money. In other words, the complex question of whether migrants are particularly susceptible to NCD deserves to be investigated, and I would argue it should be prioritised.

Migration is defined by the WHO as the “movement of a person or a group of persons, either across an international border, or within a State” (WHO, 2021). For the purpose of this literature review, a migrant is defined as anyone who resides in a country different to their country of birth, with the exception of one Indian study involving interstate migration. This study was selected due to the fact that both in terms of distance and cultural/linguistic differences, in many instances interstate migration within India is de facto akin to travelling internationally. Non-communicable disease, i.e. “chronic diseases, tend to be of long duration and are the result of a combination of genetic, physiological, environmental and behavioural factors” (WHO, 2021); these are the diseases this review will contemplate. With regards to the refugee subset of migrants, I understand the issue of confounding factors would be of particular concern, given this population is often under significant pressure in other areas, including financial strain, violence, etc.; as such they will be excluded from this study.

Accepting the broad scope of such an endeavour, the remit of the following review will focus on circumstances in a global context, given I hold that not doing so would mean not seeing the forest for the trees. I intend to research “how does migratory status affect non-communicable disease risk in a global context?” Through this question I will determine if the existing literature suffices to convey a picture on particular risk factors for NCD for migrants, and their health outcomes. I will be making research or policy recommendations in light of these findings or lack thereof.

## Methods

The aim of this review is to determine what peer-reviewed literature, particularly primary empirical research evidence has to say concerning migratory status and its effect on non-communicable disease risk in a global context, and making ad rem recommendations. A highly relevant database collection was used, one that includes relevant medical literature and collates other databases within it: EBSCOhost. The latter was used including Academic Search Complete, AMED, CINAHL and MEDLINE. These databases hold medical research and are therefore considered highly relevant for this particular question. The initial search yielded 59 results.

### Inclusion criteria

❖ Cosmopolitan literature
❖ Languages: English, Spanish, French, Portuguese, Italian
❖ Broad time frame accepted for consideration (anything published after the 1950s)
❖ Peer reviewed, empirical, primary

### Exclusion criteria

❖ Refugee-related literature
❖ Literature related to migraine

Refugee-related literature, while relevant to the concept of health, would be too complex to untangle from the concept of migration more broadly construed. Refugee populations are vastly different from migrant populations in cities, e.g. skilled migrants in Australia who are permanent residents, and as such cannot reasonably be compared to refugees in Europe through the present literature review. The search term “migra*” would yield some migraine results, but these were obviously not relevant. I find most of the issues relating to migrant health since the latter part of the 20th century are still relevant today, therefore think it prudent to include a wider range of literature than if the chosen topic had been merely communicable diseases, where treatment and prevention have drastically changed over recent decades. In any case, all selected articles are recent.

The following search string for *abstracts* was used:

- *migra\**; AND
- health; AND
- *non-communicable diseases* AND
- *risk;* NOT
- *migraine OR refugee;*

Secondary research articles were discarded; of a total of 59, about two dozen remained for abstract screening. The eight selected articles were critically appraised with the assistance of abridged CASP checklists.

The eight selected studies were the following (for a complete study characteristics table, see Appendix):

❏ Akhter, N., Begum, K., Nahar, P., Cooper, G., Vallis, D., Kasim, A. and Bentley, G.R., 2021. Risk factors for non-communicable diseases related to obesity among first-and second-generation Bangladeshi migrants living in north-east or south-east England. International Journal of Obesity, pp.1-11.
❏ Aung, T.N.N., Shirayama, Y., Moolphate, S., Lorga, T., Yuasa, M. and Nyein Aung, M., 2020. Acculturation and Its Effects on Health Risk Behaviors among Myanmar Migrant Workers: A Cross-Sectional Survey in Chiang Mai, Northern Thailand. International Journal of Environmental Research and Public Health, 17(14), p.5108.
❏ Bilal, P.I., Chan, C.K.Y. and Somerset, S.M., 2019. Acculturation and perceived ethnic discrimination predict elevated blood glucose level in Sub-Saharan African immigrants in Australia. Journal of immigrant and minority health, pp.1-7.
❏ Campostrini, S., Carrozzi, G., Severoni, S., Masocco, M. and Salmaso, S., 2019. Migrant health in Italy: a better health status difficult to maintain—country of origin and assimilation effects studied from the Italian risk factor surveillance data. Population health metrics, 17(1), pp.1-11.
❏ Eikemo, T.A., Gkiouleka, A., Rapp, C., Utvei, S.S., Huijts, T. and Stathopoulou, T., 2018. Non-communicable diseases in Greece: inequality, gender and migration. European journal of public health, 28(suppl_5), pp.38-47.
❏ Joy, E.J., Green, R., Agrawal, S., Aleksandrowicz, L., Bowen, L., Kinra, S., Macdiarmid, J.I., Haines, A. and Dangour, A.D., 2017. Dietary patterns and non-communicable disease risk in Indian adults: secondary analysis of Indian Migration Study data. Public health nutrition, 20(11), pp.1963-1972.
❏ Naicker, A., Venter, C.S., MacIntyre, U.E. and Ellis, S., 2017. Prevalence of selected intermediate risk factors for non-communicable diseases in an apparently healthy Indian community in KwaZulu-Natal, South Africa. Journal of community health, 42(1), pp.122-128.
❏ Shah, S.M., Loney, T., Sheek-Hussein, M., El Sadig, M., Al Dhaheri, S., El Barazi, I., Al Marzouqi, L., Aw, T.C. and Ali, R., 2015. Hypertension prevalence, awareness, treatment, and control, in male South Asian immigrants in the United Arab Emirates: a cross-sectional study. BMC cardiovascular disorders, 15(1), pp.1-11.

## Findings

### Critical appraisal

Eikemo et al. (2018) was focused in that it specifically targeted risk ratios and NCD rates in a specific group of migrants in Greece. The authors used a reasonable sample size (1332) that included migrants of non-Albanian origin. Their methods are somewhat convoluted by the fact that they grouped NCDs into categories and tabulated overall prevalence for several different afflictions within each category. The study is also subject to reporting bias, given that prevalence was based on self-reporting. This is particularly relevant given the study aims to study NCD reporting itself. Aung et al. (2020) focused on a more general notion of risk factors and NCDs, but again these were based on self-reported measures, not objectively measured. Their methodology seems sound, however, especially considering their simple odds ratio analysis, and Pearson correlation calculations.

Naicker et al. (2017) focused on the Indian migrant subset. Their data is particularly sound due to the fact it was clinically obtained. The “random sampling” method is not explained, however, casting a shadow of doubt over their 250 individuals’ results. At the same time, their exclusion criteria of individuals diagnosed with hypertension or diabetes mellitus is not explained. The study, however, astutely considered Asian parameter cut-offs instead of Western standards, which the authors posit more accurately reflects the Indian subpopulation. Bilal, Chan & Somerset (2019) aimed to investigate associations between acculturation, perceived ethnic discrimination (PED) and elevated blood glucose level (EBGL). Their sample size is not exceedingly large, but the clinical measurement of blood glucose was conducted appropriately. The PED questionnaire is a novel idea, but does present a level of subjectivity to the data, given that it uses a 1-5 scale for its 25 questions. This in turn poses difficulties regarding statistical analysis.

The sample used by Shah et al. (2015) consisted of 1375 individuals, with a participation rate of 76%, so drop-off rates were successfully mitigated. One of the benefits of its migrant participation was the fact that Indian, Pakistani and Bangladeshi individuals were clinically sampled, and by law all migrant workers need to undergo a communicable disease screening test, in this case encompassing TB and HIV. Their sociodemographic survey was offered in several languages, mitigating response bias. The sample used for the Italian risk factor surveillance study (Campostrini et al., 2019) was by far the largest of all the articles reviewed (over 200,000). Their investigation was restricted to country of origin, assimilation process and risk factors, making it a highly relevant focused study. With a coverage of over 90% of the Italian population, this is considered highly reliable data.

The English obesity study by Akhter et al. (2020) is highly focused, as it investigated predisposition towards obesity-related NCDs exclusively, in first and second-generation Bangladeshi migrants in England. The fact that their questionnaire was delivered face-to-face also guarantees that the data are likely to be reliable. Their study, however, might be subject to region-specific biases, as it took place in five boroughs of London and Northeastern England. The study by Joy et al. (2017) is unique in that it was the only study included in this review that represented a sample of internal migrants. I decided to include this study because of two reasons: firstly that it posed a novel question, i.e. dietary patterns and their association with micronutrient undernutrition; secondly, that as explained previously, interstate migration in India is for this review’s intents and purposes akin to international travel if one contemplates both the distance involved, and the cultural, religious and linguistic differences that abound across Indian states. The Indian Migration Study data is a treasure trove of valuable data that should not be ignored, especially considering how dietary factors seem to influence health outcomes as shown in the results section. Their sample size is quite large, and their methodology is rather simple, particularly when determining the dietary patterns themselves.

I believe that the quantitative data presented by the studies in question are generally sound, with some caveats I will discuss in the synthesis below. The information is relevant and up to date, and sources paint a coherent picture in terms of how migratory status affects non-communicable disease risk in a global context.

### Synthesis

The eight studies reviewed identify either risk factors or social determinants of health that have significant impacts on migrant health both at present and into the next decades, especially considering a post-COVID world where migration significantly ticks up. A few overarching themes have been identified, all with a quantitative backing, and will be developed as follows.

### Risk factors

Lifestyle factors among migrants were identified, wherein large percentages of the migrant population in Thailand are smokers, drinkers, obese or do not exercise (Aung et al., 2020). These are suggested to play a future role in the host country’s premature death burden. Generalisations regarding lifestyle factors are difficult to make, given a higher prevalence of smoking in South East Asia (WHO, 2021). Even though studies relied on various methodologies and took place in different continents, an emergent theme of particular determinants of health and risk factors affecting migrant populations is evoked. For instance, almost the entire sampled population surveyed by Naicker et al. (2017) were centrally obese; almost two thirds had blood pressure values above the cut-off; over a third of men and women had higher fasting blood glucose levels; and almost the entire sample recorded raised triglyceride levels. Over half of participants investigated by Shah et al. (2015) never had their blood pressure measured, and nearly a third had hypertension; notably, hypertension awareness increased with the increase in the duration of their residency for more than 5 years.

### Behaviour

Lack of exercise, alcohol consumption, smoking and central obesity were identified as major lifestyle problems (Aung et al., 2020); one must consider the harsh conditions often faced by such workers in Northern Thailand, and whether the sample was a representative one for the purposes of this review. While the important health determinant that is socioeconomic status was lower for migrants than native-born Italians (Campostrini et al., 2019), the authors also detected a lower attendance in preventive services among that population. Notably, however, this population as a whole appears not too different to native-born Italians, in terms of health attitudes and behaviours perhaps because they are mostly European migrants.

As determined by Joy et al. (2017), study participants that had increased odds of a high waist-to-hip ratio were consumers of the wheat and pulses dietary regime, while those with the greatest odds of obesity were consumers of the rice and fruit pattern, implying a strong dietary behavioural basis to NCD prevalence, at least within Indian subpopulations.

### Discrimination and integration

Albanian immigrants, particularly men could suffer increased discrimination, in turn affecting their health and their NCD reporting (Eikemo et al., 2018). Participants with a lower mean marginalisation score were more likely to smoke. 22.5% of the population studied by Bilal, Chan & Somerset. (2019) had high level PED and EBGL concomitantly. Traditional and integration modes were thus strongly correlated with EBGL and diabetes. Interestingly, PED served as a “mediating factor” between EBGL and integration mode of acculturation, a measure of how assimilated an individual is in their adopted country. Those in that mode of acculturation were 4.2 times more likely to exhibit an EBGL. Those in the traditional acculturation modes were 80% less likely to show EBGLs, which suggests a link between health and how one perceives their host country. The study’s generalisability is hampered by its small sample size and ethnic composition, however.

Migrant respondents as surveyed by Campostrini et al. (2019) seem to be “more similar” to Italians as their stay increases. The link between discrimination, both perceived and otherwise, as well as integration/acculturation warrants further study. It would also be interesting to assess whether these regional perceived discriminatory trends would apply in other regions, or are wholly European interactions.

### Gender inequality

In one of the strongest overarching data themes, gender inequalities were presented by most authors in a clear and explicit manner. To begin with, gender inequality disfavouring women in reporting NCDs both in migrants and Greek-borns was apparent. Women bear a greater risk of not reporting NCDs, particularly Albanian women, or migrants of non-Albanian origin in Greece. Risky health behaviour, as well as economic and social factors are thought to partially explain these differences in reporting (Eikemo et al., 2018). The results of Eikemo et al. (2018) suggest that gender and the occurrence of NCDs have a relationship subject to migration status as well as ethnic origin, and as such confirms that immigrants are a diverse group across social categories. Importantly, gender and migration do not represent “fixed attributes with specific health impact”, so further research is necessary to elucidate these interactions. It also begs the question of whether pregnancy influences said risks.

Being female and uneducated was associated with a lack of exercise, while alcohol consumption was associated with male Myanmarese workers (Aung et al., 2020); at the same time, central obesity was associated with uneducated females over 40 years of age in the same study. Campostrini et al. (2019) also demonstrates gender-related differences, particularly regarding attending preventive services: the prevalence is higher for Italian women and migrants. Smoking, on the other hand, is more prevalent in migrant males.

The sample surveyed by Akhter et al. (2021) showed a higher proportion of women (46% vs 10%) had a large waist circumference. According to their analysis, 42% of females had a high or very high risk of obesity-related NCDs, as opposed to 12% of males. At the same time, females had significantly higher odds of developing obesity-related NCDs, and this was particularly the case in Northeastern England. Similarly, second-generation Bangladeshis had double the odds of developing such diseases, perhaps suggesting strong acculturation, as they reflect current obesity trends in the UK. Most notably, the authors found that participants who walked over 20 minutes daily were 32% less predisposed to obesity-related NCDs (Akhter et al., 2021). The article by Joy et al. (2017) also exposed gender-related patterns, including an inadequate calcium intake in 13% and 42% of females in the rice and fruit, and rice & low diversity dietary patterns respectively.

All in all, findings presented by the eight selected articles provide insight into the research question. Considering the question was restricted to a global context yet most research centred on individual communities, there is ample possibility for further study concerning risk factors and NCD association analysis in a collation of migrants. This would particularly call for an international meta-analysis approach.

## Discussion and recommendations

### Key findings

Peer-reviewed, primary empirical research evidence tells us the following about non-communicable disease risk in a global context:

❏ Migrant populations are heterogeneous
❏ Certain migrant subgroups show increased risk of developing NCDs in some circumstances
❏ Behaviour plays a crucial role in how these risks are distributed
❏ Perceived discrimination is a novel factor to consider when establishing risk associations
❏ Gender-related differences exist in most migrant populations
❏ Gaps in the research exist

### In context

Identifying and quantifying risk factors for migrants is a vital aspect of health science and requires continuous investigation and monitoring. This review identifies several such risk factors, namely: sex, smoker status, obesity, PED, integration, alcohol consumption, blood pressure, blood glucose level, and notably one protection factor: walking 20 minutes or more daily.

While research usually focuses on risk factors, protective factors are just as important, and research such as that undertaken by Akhter et al. (2021) was refreshing in that it considered such a notion. One notable absence from this particular set of articles was the issue of refugees, and the effect this circumstance has on both risk factors and health outcomes. This was a consequence of my selection criteria, but it should be noted that it likely would be relevant for a meta-analysis. The findings of this review are important in that they provide fertile ground for comparison and appropriate planning, as some of the conclusions might be applicable in different countries or localities, at any age, and with particular importance given to gender disparities. Risk factors can easily be incorporated into modelling and subsequently applied as part of public health measures, such as Smit et al. (2017) and Brinks & Landwehr (2014) suggested, albeit in a slightly different milieu. Finally, as suggested by Aung et al. (2020), these factors could create a wave of premature deaths in the host country, with wide ranging future implications for all involved parties.

### Recommendations

As a direct outcome of this review, identifying a handful of the most powerful risk factors such as obesity, smoking status, sex, and protective factors such as whether one walks for 20 minutes or more daily can very quickly be applied in policy, e.g. through a widespread advertisement campaign. Public health initiatives should be simple to implement based on the observations collated by this review. Concomitantly, preventive healthcare checks are an extremely valuable tool, particularly in primary care conditions, and are ones that migrant populations are particularly prone to ignore, as previously discussed. At an international level, cooperative efforts in planning can be strongly influenced by this research, such as the deployment of international health passports or an electronic medical history that can be translated to any language on demand. Another prospect would be the global standardisation of preventive health tests for different ages: blood pressure for at risk adult populations, pregnancy controls for women, eyesight tests for vulnerable youths, etc. As long as migration takes place, and in fact increases, such research will continue to be justified.

Research on protective factors is both relevant and highly recommended, as one of the studies reviewed demonstrated. Changes to populational patterns such as those demanded of obese or alcoholic individuals are extremely hard to achieve, nevertheless efforts can be targeted and treatments can be better directed to such in-risk population strata, before and after they have migrated in order to diminish the public health burden accepted when allowing migration to occur. Barriers in funding can also be significant, especially considering countries with a strong private healthcare sector which routinely sets an economical barrier for migrants. This needs to be addressed by governments as a matter of urgency, as even small educational campaigns or free testing can go a long way in demystifying diseases and their prevention.

I suggest the following research focuses in light of the review: meta-analyses that also account for refugee status; studies into the effects of obesity pathophysiology in migrant populations globally; studies into migrant social and gender inequalities vis à vis NCDs; further research into socioeconomic status and its effect on a specified set of risks, including those related to diet and sex; studies about NCD protective factors.

In more practical terms, it would be particularly relevant for future research to investigate whether migrant women are particularly prone to obesity-related NCDs after giving birth, in a way combining Bilal, Chan & Somerset’s (2019) methodology with the sample reliability exhibited by Campostrini et al. (2019).

### Strengths and limitations

This review’s strengths were two-fold. First, its specific scope made selection criteria stringent, and therefore the selected papers had already undergone heavy scrutiny. Available research was rather extensive, albeit of mixed relevance. The articles screened had valuable information which provided data particularly relevant for this review. The studies being quantitative and with relatively large sample sizes in most cases meant that their results were largely reliable and generalisations within a global context could prudently be allowed in some instances.

At the same time, this review included literature from a global perspective, including studies undertaken in India, England, Australia, the UAE, etc. This could be interpreted as both a strength and a weakness concomitantly: it is important to identify global trends regarding NCDs in the migrant population, but these trends could be difficult to identify if “watered down” by contradicting data from disparate locations. For instance, a risk factor relevant for South Asian migrants in the UAE might not necessarily be relevant for Sudanese migrants in Melbourne.

The most pressing weakness could arguably be the fact that refugee-related results were excluded from the review. These are some of the most vulnerable migrant subsets, and as such deserve focused research that takes into account the complexities associated with such a circumstance, which in and of itself is dynamic and depends on a myriad factors outside this review’s purview, and would better suit a meta-analysis.

## Data Availability

All data pertaining to the study was extracted from the articles reviewed.

## Appendix

**Figure 1.**
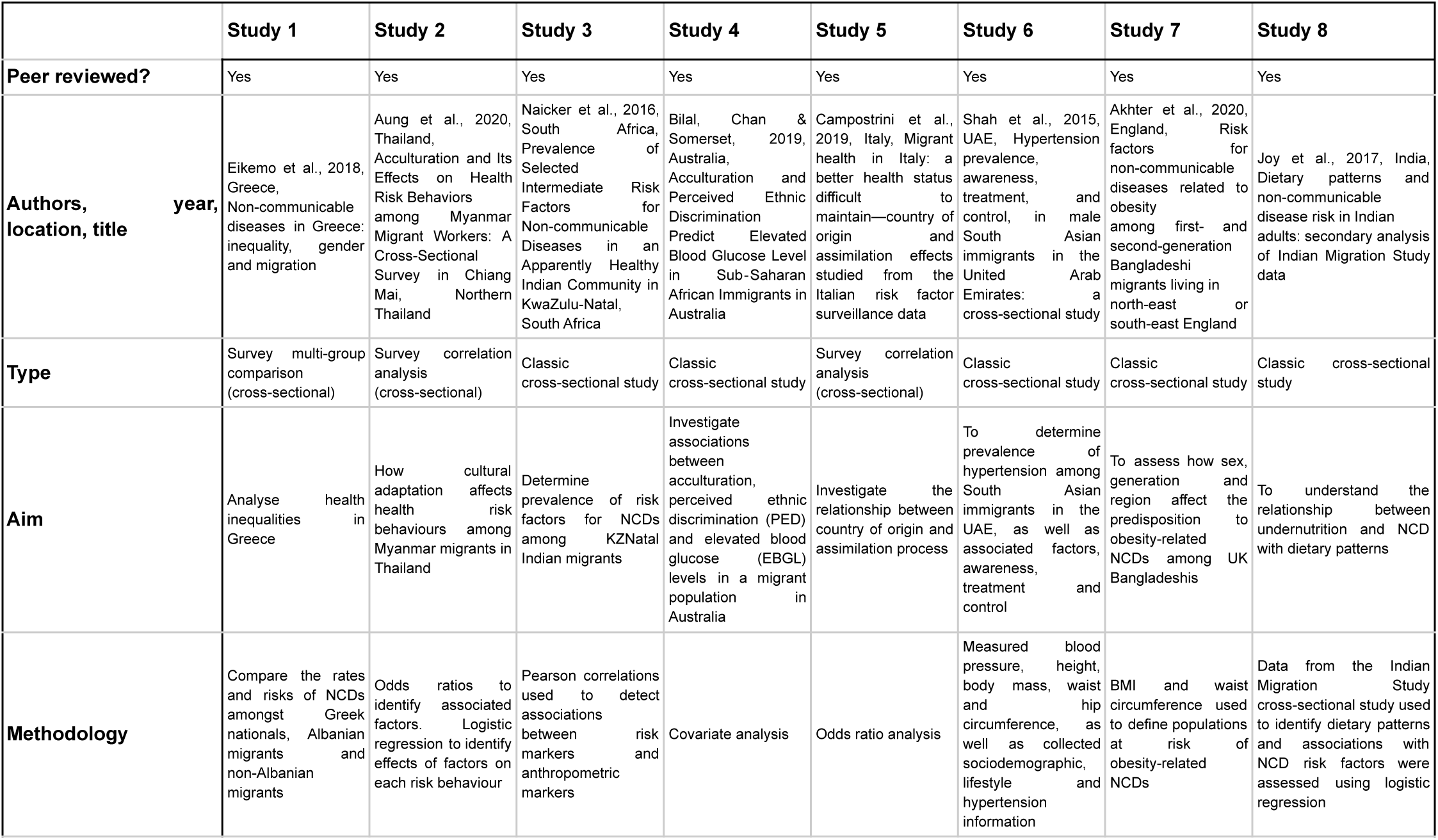
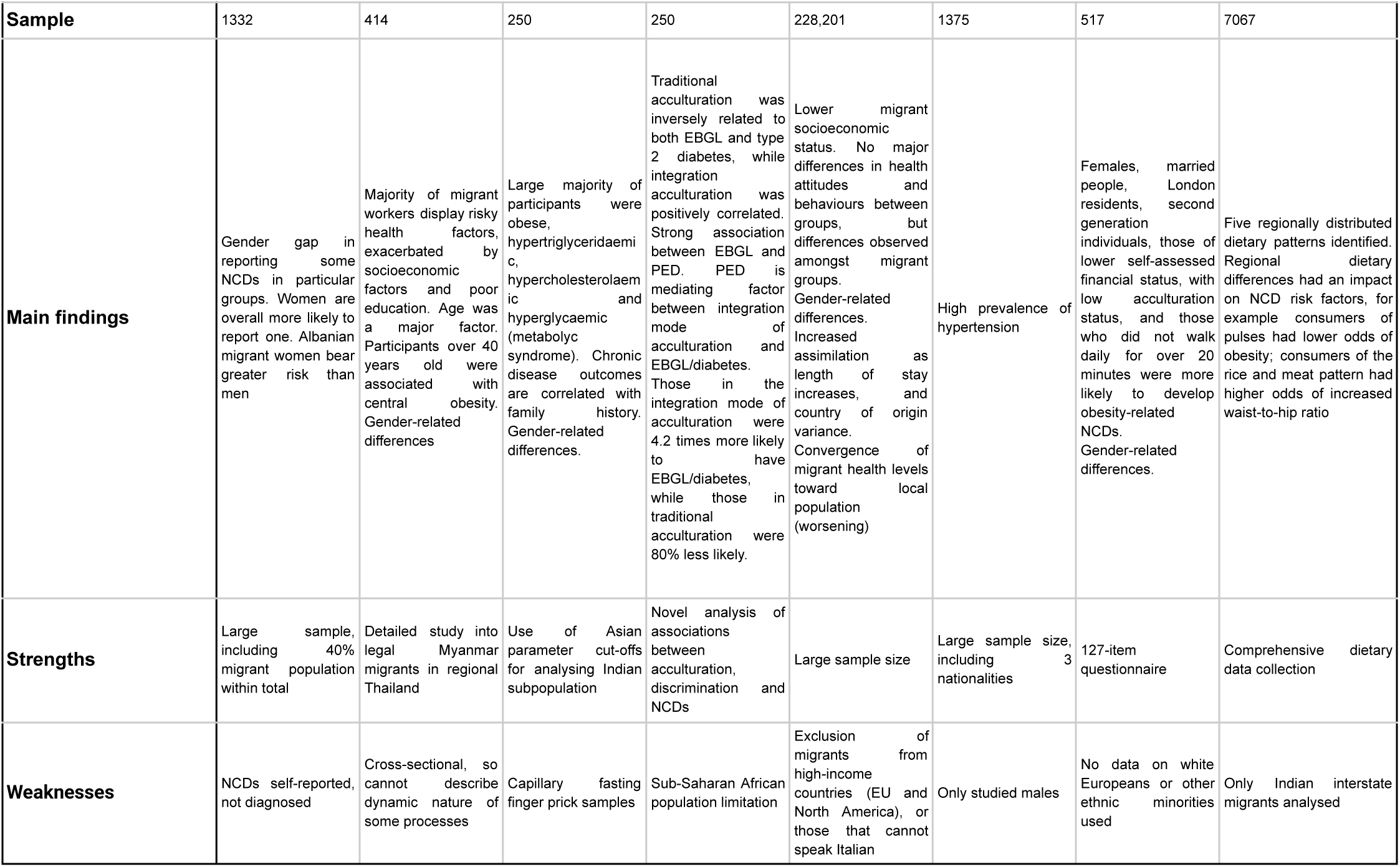
Study characteristics table including the 8 reviewed articles.

## Notes

### Competing Interest Statement

The authors have declared no competing interest.

### Funding Statement

No funding was obtained to finance this study.

